# Classification of Primary Angle Closure Disease by Hierarchical Cluster Analysis of Ocular Biometrics in the Dark and Light

**DOI:** 10.1101/2023.01.18.23284636

**Authors:** Austin Cho, Juan Pablo Lewinger, Anmol A. Pardeshi, Galo Apolo Aroca, Mina Torres, Monisha Nongpiur, Xuejuan Jiang, Roberta McKean-Cowdin, Rohit Varma, Benjamin Y. Xu

**Affiliations:** Keck School of Medicine of the University of California, Los Angeles, CA; Roski Eye Institute, Keck School of Medicine, University of Southern California, Los Angeles, CA; Department of Population and Public Health Science, Keck School oof Medicine at the University of Southern California, Los Angeles, CA; Southern California Eye Institute, CHA Hollywood Presbyterian Medical Center, Los Angeles, CA; Singapore Eye Research Institute and Singapore National Eye Center, Singapore, Republic of Singapore

**Author notes:** **Corresponding Author:** Benjamin Xu, Department of Ophthalmology, Keck School of Medicine at the University of Southern California, 1450 San Pablo Street, 4th Floor, Suite 4700, Los Angeles, CA 90033.

## Abstract

**Purpose:** To investigate the classification of eyes with primary angle closure disease (PACD) based on hierarchical cluster analysis of ocular biometrics measured in the dark and light using anterior segment OCT (AS-OCT).

**Methods:** Participants of the Chinese American Study received complete eye exams, including gonioscopy and AS-OCT imaging, to identify primary angle closure suspects (PACS) and primary angle closure without/with glaucoma (PAC/G). Biometric parameters analyzed included angle opening distance (AOD750) and trabecular iris space area (TISA750), iris area (IA), iris thickness (IT750), iris curvature (IC), lens vault (LV), anterior chamber width (ACW) and anterior chamber depth (ACD). Hierarchical cluster analysis was performed using Ward’s method and Euclidean distance.

**Results:** Analysis of 159 eyes with PACS or PAC/G produced 2 clusters in both dark and light. In both analyses, the primary cluster (N=132 in the dark, N=126 in the light) was characterized by smaller AOD750 and TISA750, greater IC, and greater LV (p<0.001). The proportion of PACS to PAC/PACG eyes was significantly different between clusters in the light (p=0.02) but not the dark cluster analysis (p=0.08). On multivariable logistic regression analysis, smaller TISA750 (OR=0.84 per 0.01μm^2^) and AOD750 (OR=0.93 per 0.01mm) in the light and smaller TISA750 (OR=0.86 per 0.01μm^2^) in the dark were significantly associated (p≤ 0.02) with higher odds of PAC/G.

**Conclusion:** Cluster analysis of ocular biometrics can classify PACD eyes by disease severity. Ocular biometrics appear equally if not more strongly predictive of disease severity when measured in the light than dark. Clustering of biometric measurements obtained in the light could provide a novel method to risk-stratify patients for more severe PACD.

## Introduction

Primary angle closure glaucoma (PACG) is a leading cause of blindness worldwide.^1,2^ Primary angle closure disease (PACD) is a spectrum of disease characterized by appositional or synechial closure of the anterior chamber angle.^3^ Angle closure in the form of iridotrabecular contact can impede aqueous flow through the trabecular meshwork, progressing to higher intraocular pressure (IOP) and, in severe cases, PACG.^4^ Angle closure eyes are typically categorized as primary angle closure suspects (PACS), primary angle closure (PAC), and primary angle closure glaucoma (PACG) across this spectrum of disease severity.^5^

Ocular biometric parameters measured using anterior segment OCT (AS-OCT) are well-established risk factors in PACD.^6–10^ These parameters can also provide information about underlying anatomical mechanisms of angle closure, such as pupillary block, plateau iris configuration, exaggerated lens vault, or thick peripheral iris.^7, 11–13^ For example, greater iris curvature is believed to reflect increased pupillary block as aqueous humor collects posterior to the iris, and iris thickness, which can be directly measured in AS-OCT images, directly contributes to higher risk of angle closure.^14,15^ Hierarchical cluster analysis, an unsupervised analysis method, also identifies patterns among ocular biometric measurements that appear to conform to these broad categories of angle closure mechanisms.^16–19^

Clinical assessments of angle closure risk and underlying anatomical mechanisms are by convention conducted in the dark as angle width tends to be narrower in the dark than in the light.^20^ However, despite strong associations between ocular biometric measurements and presence of PACD, static measurements under dark lighting conditions appear only moderately predictive of disease severity and progression.^21,22^ This raises the question whether biometric data obtained under different lighting conditions might provide additional information about PACG risk, especially since the majority of waking hours are typically spent in lit environments. While more recent studies have begun to assess anatomical differences between the dark and light, the impact of these differences on anatomical mechanisms and disease severity remains unclear.^20,23^

In this study, we perform unsupervised hierarchical cluster analysis of biometric data from the Chinese American Eye Study (CHES) to classify eyes with mild (PACS) and severe (PAC and PACG) PACD. The CHES data comprises the full spectrum of PACD severity in contrast to prior cluster analysis studies of angle closure eyes.^16–18^ In addition, we analyze biometric data obtained in the dark and light to identify differences in clustering patterns of ocular biometric parameters and PACD severity. While there is limited knowledge about the clinical significance of biometric measurements obtained in the light, we hypothesize that clustering patterns may differ under the two lighting conditions, which could provide novel insights into disease mechanisms and severity.

## Methods

The Chinese American Study (CHES) was approved by the Ethics Committee from the University of Southern California Medical Center Institutional Review Board. All procedures followed the recommendations of the Declaration of Helsinki. All participants gave informed consent at the time of enrollment. CHES study participants were recruited as part of a population-based study on ocular disease in Chinese American individuals aged ≥ 50 years living in Monterey Park, California. ^20^ Patients with any history of eye procedures, including laser peripheral iridotomy and cataract surgery, that could affect the anterior segment structures were excluded from this study.

### Clinical Examination

Each participant received a complete eye examination by a trained ophthalmologist, including manual gonioscopy and AS-OCT imaging (CASIA SS-1000; Tomey Corporation) in the upright seated position. Gonioscopy was performed under dark ambient lighting (0.1 candela [cd]/m^2^) with a 1-mm light beam and a Posner-typer 4-mirror lens (Model ODPSG; Ocular Instruments, Inc) by a trained ophthalmologist (D.W.). The angle in each quadrant was graded according to the modified Shaffer classification system: grade 0, no structures visible; grade 1, nonpigmented TM visible; grade 2; pigmented TM visible; grade 3, scleral spur visible; and grade 4, ciliary body visible. PACD was defined as an eye with ≥ 3 quadrants gonioscopic angle closure (grade 0 or 1) in the absence of potential causes of secondary angle closure such as inflammation or neovascularization.

Primary angle closure suspect (PACS) was defined as narrow angles with IOP ≤ 21mmHg without peripheral anterior synechiae (PAS). Primary angle closure (PAC) was defined as PACS with IOP > 21 mmHg or PAS without evidence of glaucomatous optic neuropathy (GON). PACG was defined as PAC with evidence of GON. PAC and PACG were grouped together as PAC/G in this study due to the relatively small number of PACG cases in the CHES cohort.

### Anterior Segment Optical Coherence Tomography

AS-OCT imaging of both eyes was performed under dark (0.1 cd/m^2^) and light (27 cd/m^2^) ambient lighting conditions before pupillary dilation. The SS OCT viewer software (V.3.0) was used to automatically segment structures and measure anterior segment biometric parameters after an experienced grader (A.P.) manually identified the scleral spur (SS) in each image. The grader was masked to the identities and other exam findings of the participants. One eye per participant was selected at random for analysis using MATLAB software (MathWorks, Natick, MA). Four of 128 two-dimensional radial cross-sectional images were analyzed per eye, measuring up to 8 different sectors. Sectoral measurements were then averaged, and the average measurement value was used for this study. Eyes missing measurements in 4 or more sectors were excluded from the analysis.

Two biometric parameters describing angle width were measured: angle opening distance (AOD) and trabecular iris space area (TISA).^24^ AOD750 was defined as the perpendicular distance from the TM at 750 μm anterior to the scleral spur to the anterior iris surface. TISA750 was defined as the area bounded anteriorly by AOD750; posteriorly by a line drawn from the scleral spur perpendicular to the plane of the inner scleral wall to the opposing iris; superiorly by the inner corneoscleral wall; and inferiorly by the iris surface. Iris area (IA), anterior chamber depth (ACD), iris curvature (IC), lens vault (LV), and anterior chamber width (ACW) were also measured.^24,25^ IA was defined as the cross-sectional area of the full length of the iris. ACD was defined as the distance from the apex of the anterior lens surface to the apex of the corneal endothelium. IC was defined as the distance from the apex of the iris convexity to a line extending from the peripheral to central iris pigment epithelium. ACW was defined as the distance between scleral spurs. Pupil diameter (PD) was defined as the shortest distance between the edges of the pupil. Intra-grader measurement repeatability was previously assessed and reported to be excellent for all parameters, with intraclass correlation coefficients (ICCs) ranging from 0.89 to 0.98.^20^

### Statistical Analysis

Continuous and categorical variables were summarized as mean ± standard deviation (SD) and proportions, respectively. Distributions of continuous variables were compared using the two-sample t-test or Wilcoxon signed rank test depending on the result of normality testing using the Shapiro-Wilk test. Proportions of categorical variables were compared using the Chi-squared test. Hierarchical cluster analysis was used to classify PACD eyes. Measurements of each parameter were standardized (values subtracted by mean and then divided by standard deviation) prior to analysis so as not to affect squared Euclidean distances. Agglomerative cluster analysis was performed using Euclidean distance as the similarity measure and Ward’s method as the clustering algorithm. Each case started with each cluster as a separate cluster; clusters were then combined until only one cluster remained. Cluster analysis was applied used squared Euclidean distances as a similarity measure and Ward’s method as the clustering algorithm. The Duda-Hart (DH) index and pseudo t^2^ statistics (PST2) were used to determine the optimal number of clusters. The DH index utilizes the ratio of the two within sum of squares to decide if a cluster can be divided into separate clusters. PST2 is derived from the DH index and accounts for the total number of cases. The optimal number of clusters was chosen by selecting the number of clusters corresponding to peaks in the PST2 value and a low DH index value. Univariable logistic regression analysis was performed with dark and light AS-OCT parameters as independent variables and disease severity (PACS or PAC/PACG) as the dichotomous outcome variable. Multivariable logistic regression analysis included age, sex, and all parameters with p-value < 0.2 on univariable analysis; AOD750 and TISA750 were not included in the same model due to collinearity. Area under the receiver operating curve (AUC) metrics were calculated to assess predictive performance of regression models. All analyses were performing using the R software version 4.2.1 (R Foundation for Statistical Computing, Vienna, Austria). Statistical Analysis were conducted using a significance level of 0.05.

## Results

AS-OCT data was available on 169 eyes of 169 participants with PACS or PAC/G. After excluding 10 patients with history of intraocular surgery or LPI, 159 eyes were eligible for analysis, all of which had biometric data from 5 or more sectors. 120 eyes had PACS and 39 participants had PAC/G. Mean age was 61.7 ± 7.79, mean IOP was 16.2 ± 3.49, and 122 (76.7%) of the participants were female. The optimal number of clusters was 2 for both dark and light measurements based on DH index and PST2.

In the dark analysis, there were 132 eyes in Cluster 1 and 27 eyes in Cluster 2 **(Table 1, Figure 1)**. Cluster 1 was significantly (p = 0.03) older than Cluster 2 (62.3 ± 7.9 and 58.8 ± 6.8 years, respectively). Clusters 1 and 2 had similar IOP (16.34 ± 3.6 and 15.3 ± 2.7 mmHg, respectively; p = 0.14) and proportion of females overall (75.8% and 81.4%, respectively; p = 0.52). Cluster 1 had smaller AOD750 and TISA750 and greater IC and LV (p < 0.001) **(Table 1)**. Cluster 2 had greater ACD and PD (p < 0.001). There was no significant difference (p = 0.12) in the proportion of PAC/G between Clusters 1 (36 out of 132; 27.3%) and 2 (3 out of 27; 11.1%) in the dark analysis.

**Table 1.** Comparison of demographics and ocular biometric factors between Clusters 1 and 2 from the 2-cluster analysis.

| Lighting | Mean Parameters (Mean<br>± SD) | Cluster 1 | Cluster 2 | P-value<sup>a</sup> |
| --- | --- | --- | --- | --- |
| Dark |  | N = 132 | N = 27 |  |
|  | Sex (F:M)<sup>b</sup> | 100:32 | 22:5 | 0.52 |
|  | Age (years)<sup>b</sup> | 62.3 ± 7.87 | 58.8 ± 6.78 | **0.03** |
|  | IOP<sup>b</sup> (mmHg) | 16.368 ± 3.616 | 15.321 ± 2.688 | 0.14 |
|  | PAC/G:PACS<sup>c</sup> | 36:96 | 3:24 | 0.08 |
|  | AOD750 (mm) | 0.157 ± 0.051 | 0.225 ± 0.055 | < **0.001** |
|  | TISA750 (mm<sup>2</sup>) | 0.088 ± 0.034 | 0.114 ± 0.024 | < **0.001** |
|  | IA (mm<sup>2</sup>) | 1.611 ± 0.213 | 1.554 ± 0.213 | 0.24 |
|  | IT750 (mm) | 0.398 ± 0.080 | 0.425 ± 0.052 | **0.02** |
|  | IC (mm) | 0.297 ± 0.060 | 0.206 ± 0.060 | < **0.001** |
|  | ACD (mm) | 2.137 ± 0.195 | 2.420 ± 0.184 | < **0.001** |
|  | LV (mm) | 0.838 ± 0.157 | 0.573 ± 0.136 | < **0.001** |
|  | ACW (mm) | 11.509 ± 0.195 | 11.454 ± 0.281 | 0.49 |
|  | PD (mm) | 3.670 ± 0.657 | 4.259 ± 0.880 | < **0.001** |
| Light |  | N = 126 | N = 33 |  |
|  | Sex<sup>b</sup> (F:M) | 98:28 | 24:9 | 0.54 |
|  | Age<sup>b</sup> (years) | 62.1 ± 8.26 | 60.5 ± 5.61 | 0.66 |
|  | IOP<sup>b</sup> (mmHg) | 16.306 ± 3.557 | 15.747 ± 3.236 | 0.36 |
|  | PAC/G:PACS<sup>c</sup> | 36:90 | 3:30 | **0.02** |
|  | AOD750 (mm) | 0.218 ± 0.054 | 0.311 ± 0.047 | < **0.001** |
|  | TISA750 (mm<sup>2</sup>) | 0.126 ± 0.034 | 0.164 ± 0.026 | < **0.001** |
|  | IA (mm<sup>2</sup>) | 1.811 ± 0.221 | 1.888 ± 0.218 | **0.08** |
|  | IT750 (mm) | 0.339 ± 0.065 | 0.332 ± 0.046 | 0.58 |
|  | IC (mm) | 0.301 ± 0.072 | 0.256 ± 0.056 | < **0.001** |
|  | ACD (mm) | 2.101 ± 0.168 | 2.464 ± 0.142 | < **0.001** |
|  | LV (mm) | 0.867 ± 0.158 | 0.656 ± 0.124 | < **0.001** |
|  | ACW (mm) | 11.483 ± 0.375 | 11.685 ± 0.362 | **0.006** |
|  | PD (mm) | 2.553 ± 0.592 | 2.567 ± 0.351 | 0.32 |

ACD: Anterior chamber depth; ACW: Anterior chamber width; AOD: Angle opening distance; IA: Iris area; IC: Iris curvature; IOP: Intraocular pressure; IT: Iris thickness; LV: Lens vault; PAC: Primary angle closure; PACS: Primary angle closure suspect; PACG: Primary angle closure glaucoma; PD: Pupillary diameter; TISA: Trabecular iris surface area
<sup>a</sup> Statistical significance tested by Wilcoxon t-test or two-sample t-test
<sup>b</sup> Post hoc; not included in cluster analysis
<sup>c</sup> Statistical Significance tested by Chi-Squared test

**Figure 1:**
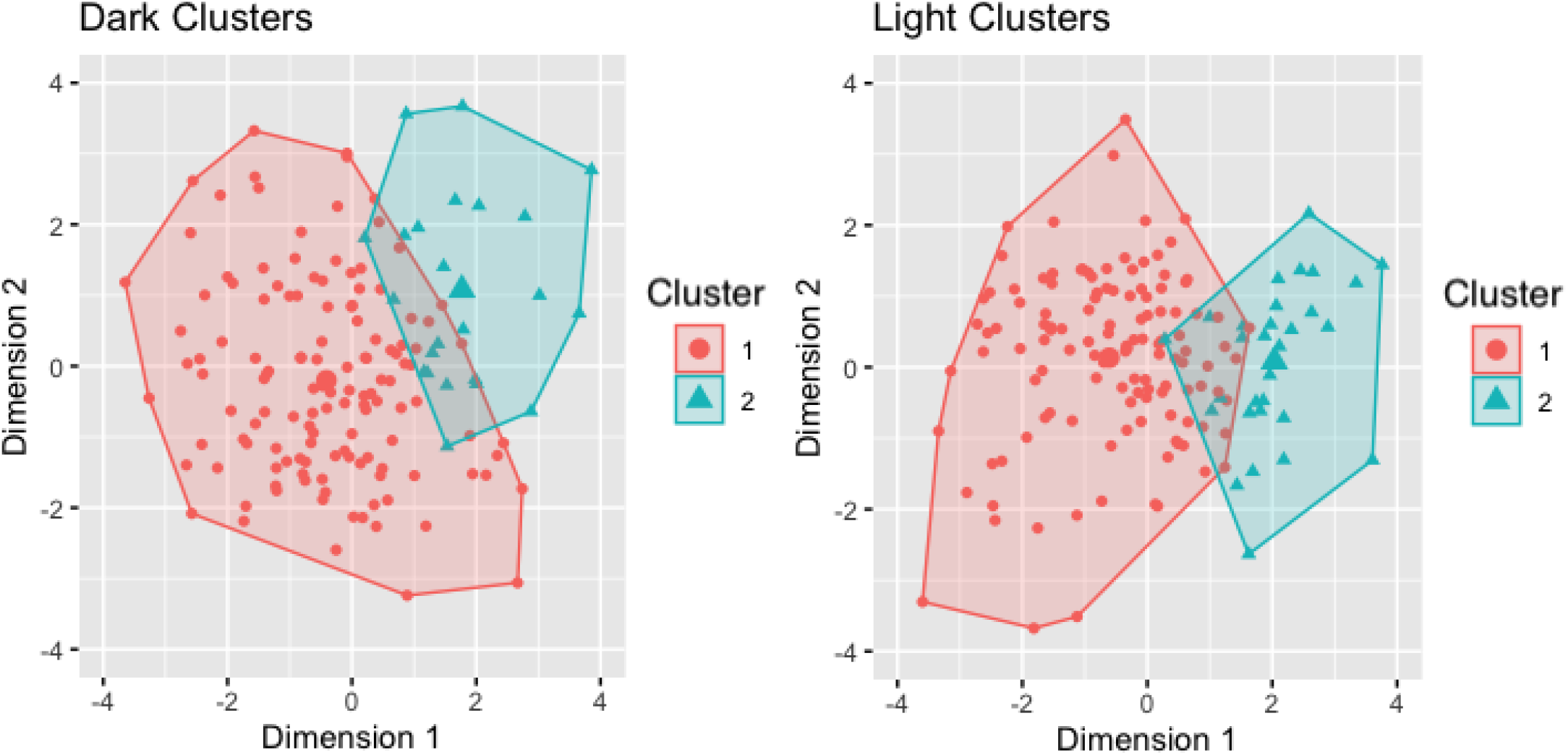
Clustering of AS-OCT measurements from PACD eyes in the dark (left) and light (left).

In the light analysis, cluster 1 had 126 eyes and cluster 2 had 33 eyes **(Table 1, Figure 1)**. No significant difference was found (p = 0.66) in age between Cluster 1 and Cluster 2 (62.1 ± 8.3 and 60.5 ± 5.6 years, respectively). Clusters 1 and 2 had similar IOP (16.3 ± 3.6 and 15.7 ± 3.2, respectively; p = 0.36) and proportion of females overall (77.8% and 72.7%; p = 0.54). Cluster 1 had smaller AOD750 and TISA (p < 0.001), smaller ACW (p = 0.01), and greater IC and LV (p < 0.001) **(Table 1)**. Cluster 2 had greater ACD (p < 0.001). There was a significantly greater (p = 0.04) proportion of PAC/G in Cluster 1 (36 out of 126; 28.6%) compared to Cluster 2 (3 out 33; 9.1%) in the light analysis.

In an analysis of change in cluster between dark and light, cluster identity was mostly conserved across lighting conditions **(Table 2)**. 13 eyes (8.2% of total; 11 PACS, 2 PAC/G) changed from Cluster 1 in the dark analysis to Cluster 2 in the light analysis. 7 eyes (4.4% of total; 5 PACS, 2 PAC/G) changed from Cluster 2 in the dark analysis to Cluster 1 in the light analysis.

**Table 2.** Number of eyes that switched clusters between dark and light analyses.

| Clusters | PACS | PAC/G | Total # |
| --- | --- | --- | --- |
| Cluster 1 only (dark and light) | 85 | 34 | 119 |
| Cluster 2 only (dark and light) | 19 | 1 | 20 |
| Cluster 1 (dark) to Cluster 2 (light) | 11 | 2 | 13 |
| Cluster 2 (dark) to Cluster 1 (light) | 5 | 2 | **7** |
| PAC: Primary angle closure; PACS: Primary angle closure suspect; PACG: Primary angle closure glaucoma |  |  |  |
| <sup>a</sup>Dark Analysis |  |  |  |
| <sup>b</sup>Light Analysis |  |  |  |

A sensitivity analysis conducted with 3 instead of 2 clusters produced similar results, except Cluster 1 was sub-divided into 2 clusters, Clusters 1A and 1B **(Supplementary Table 1, Supplementary Figure 1)**. In the dark analysis, Cluster 1A (71) was characterized by smaller AOD750 and TISA750; Cluster 1B (61) was characterized by greater IC and LV; Cluster 2 (27) was characterized by greater ACD, IT, and PD. In the light analysis, Cluster 1A (110) was characterized by greater IC; Cluster 1B (16) was characterized by smaller AOD750 and TISA750, ACW, and PD; Cluster 2 (33) was characterized by greater ACD. Similar to the 2-cluster analysis, there was a significant inter-cluster difference in proportion of PAC/G in the light analysis (p = 0.01), but not in the dark analysis (p = 0.07).

Post hoc analysis was performed to identify factors contributing to the difference in proportion of PAC/G between clusters. On univariable logistic regression analysis, only greater TISA750 in the dark (OR = 0.86 per 0.01μm^2^) and light (OR = 0.84 per 0.01μm^2^) and greater AOD750 in the light (OR = 0.93 per 0.01μm) were significantly associated (p = 0.02) with lower odds of PAC/G **(Table 3)**. On multivariable logistic regression analysis of eligible parameters measured in the light, greater TISA750 (OR = 0.85 per 0.01 µm^2^) was significantly associated (p = 0.007) with lower odds of PAC/G after adjusting for age and sex (area under the receiver operating characteristic curve [AUC] = 0.681, 95% CI: 0.573-0.788). Greater AOD750 (OR = 0.93 per 0.01μm) in the light was also significantly associated (p = 0.04) with lower odds of PAC/G in a separate multivariable model with similar covariates (AUC = 0.641, 95% CI: 0.538 – 0.744). TISA750 was the only parameter measured in the dark eligible for multivariable logistic regression analysis; greater TISA750 (OR = 0.85 0.01 per µm^2^) remained associated (p = 0.01) with lower odds of progression after adjusting for age and sex (AUC = 0.628, 95% CI: 0.525 – 0.782).

**Table 3.**
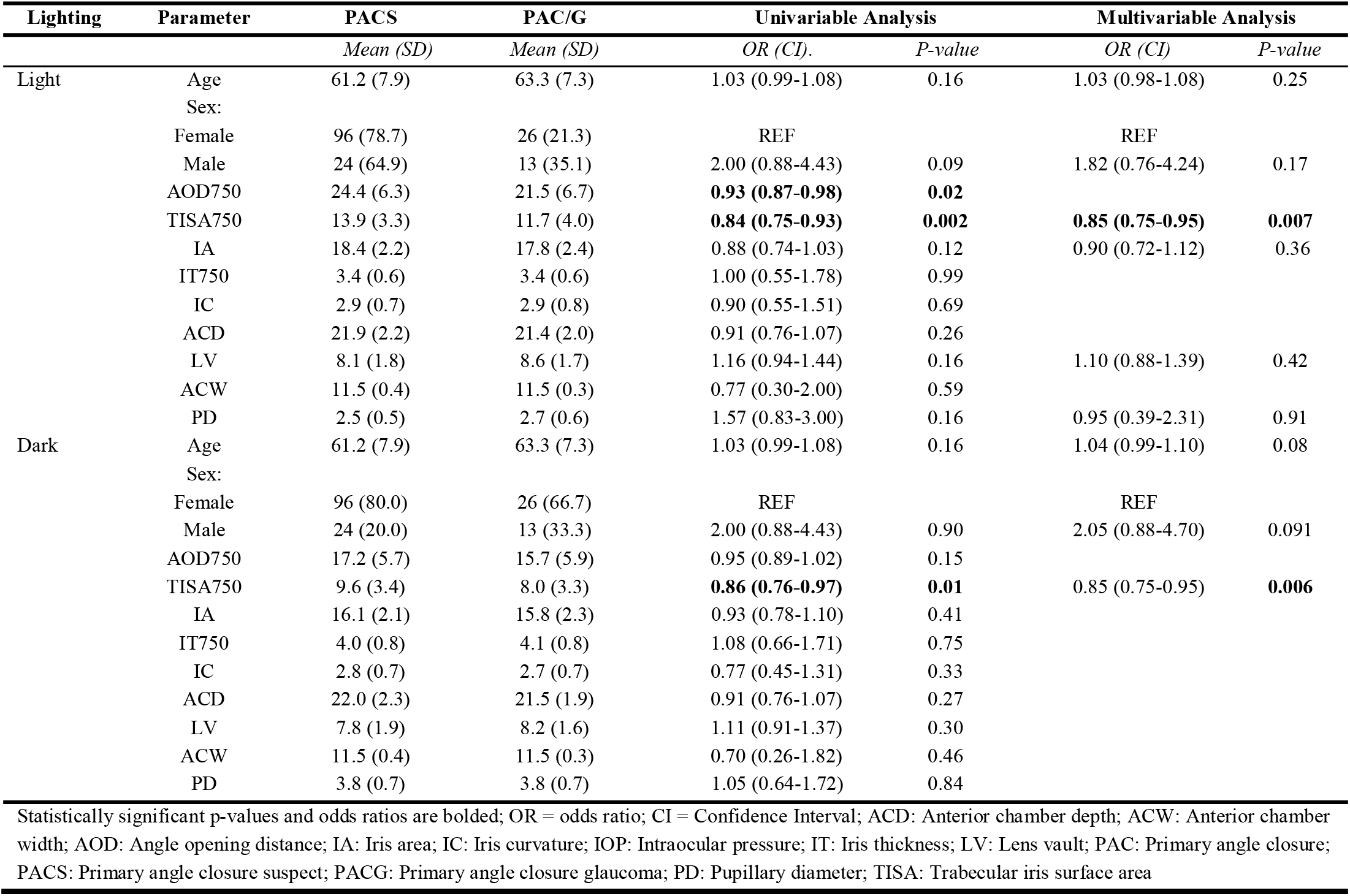
Univariable and multivariable logistic regression analysis of demographic and ocular biometric factors associated with PAC/G.

## Discussion

In this study, hierarchical cluster analysis of biometric measurements from Chinese American eyes with PACD revealed two clusters under both light and dark conditions. Clusters in dark and light analyses both appeared to segregate based primarily on established biometric risk factors for PACD. While Cluster 1 had a significantly higher proportion of severe PACD (PAC/G) than Cluster 2 in the light, inter-cluster difference in proportions was not significantly different in the dark. Post hoc logistic regression analysis showed that smaller TISA750 and AOD750 in the light and TISA750 in the dark were significantly associated with disease severity. These findings based on unsupervised analysis of PACD eyes provide potential insights into disease mechanisms and the role of biometric measurements obtained in the light for risk stratifying patients for more severe PACD.

Our cluster analysis produced 2 clusters in both the light and dark analyses, similar to cluster analyses of PACD eyes conducted by Baek et al. and Nongpiur et al.^16,18^ Similar to in those studies, clusters in our study did not appear cleanly grouped by angle closure subtype (e.g. pupillary block, plateau iris configuration, thick peripheral iris roll). Instead, biometric factors that are associated with more severe PACD were generally grouped together.^16,18^ Cluster 1 in both dark and light analyses were characterized by narrower angles, greater IC and LV, and smaller ACD, which are well-established biometric risk factors for PACD.^6,7,9^ It is tempting to further sub-categorize the clusters into angle closure subtypes; increased IC has been previously reported as an indicator of pupillary block, and flatter IC and deeper ACD are associated with plateau iris configuration.^26,27,28^ However, this type of sub-classification should be performed cautiously as angle closure subtypes cannot be confirmed with existing CHES data (e.g. UBM was not performed to confirm plateau iris configuration), and it is difficult to ascertain why the unsupervised cluster analysis grouped eyes together in the way it did. Therefore, we believe it is only appropriate to conclude that cluster analysis appears to segregate PACD eyes in CHES by factors that are known to increase risk of angle closure.

Cluster analysis of biometric measurements in CHES appeared to identify a sub-population of eyes with a higher proportion of severe PACD (Cluster 1). This finding highlights a key difference between our study, which includes PACS and PAC/G eyes, and studies by Baek et al., which only included PAC/G patients, and Nongpiur et al., which only included either PACS or PAC/G eyes in two separate cluster analyses.^16–18^ Specifically, when eyes that span the entire spectrum of PACD are clustered together, classification appears to occur by disease severity. Our cohort also differs from that of Moghimi et al., which included a substantial proportion (nearly half) of APAC and fellow APAC eyes.^19^ This difference may explain why their analysis produced 3 clusters, with one cluster comprised almost entirely of APAC and fellow APAC eyes. These differences in results between studies suggest that this type of clustering analysis, and by extension inferences about its results, may be specific to the study cohort.

The identification of a sub-population of eyes with a higher proportion of severe PACD is clinically significant in the setting of the Zhongshan Angle-Closure Prevention (ZAP) Trial and Singapore Asymptomatic Narrow Angles Laser Iridotomy Study (ANA-LIS), which both reported low risk of progression from PACS to PAC or acute angle closure (AAC).^29,30^ These studies highlight the importance of developing novel methods to risk-stratify PACS eyes for more severe disease and identify eyes that could benefit from prophylactic treatment with laser peripheral iridotomy (LPI). While is tempting to speculate that PACS eyes in Cluster 2 may be at higher risk of developing PAC/G than in Cluster 1, it is important to point out that our findings are based on cross-sectional data that could be confounded by unobserved factors. Therefore, cluster analysis of data from longitudinal studies like the ZAP Trial and ANA-LIS may help establish the relative prognostic value of clustering biometric measurements for predicting angle closure progression starting from a common baseline.

Cluster analysis of biometric measurements from the light but not the dark segregated eyes into groups with significantly higher (Cluster 1; 29% probability) or lower (Cluster 2, 9% probability) proportion of eyes with PAC/G. While the measurements of most biometric parameters differed between Clusters 1 and 2 in the light, it is unclear which parameters contributed to the difference in disease severity based solely on the unsupervised cluster analysis. This led us to perform post hoc logistic regression analysis of biometric measurements as risk factors for PAC/G, which showed that angle width parameters were the only significant predictors of disease severity, consistent with findings by Moghimi et al.^31^ This finding is also consistent with a previous study by Xu et al. that found smaller angle width predicted progression from PACS to PAC or AAC in ZAP Trial participants.^6^ Comparison of biometric measurements from Clusters 1 and 2 in the dark showed some of the same differences as in the light; however, the proportion with PACG (27% and 13%, respectively) was not significantly different; although, this may be an effect of the relatively small sample size of PAC/G eyes in our cohort. In addition, only TISA750 and not AOD750 was associated with disease severity in the dark. Finally, the AUC of TISA750 trended toward being higher in the light than the dark. These findings together suggest that angle width is an important parameter when evaluating eyes for disease severity in both the light and dark. Furthermore, angle parameters are equally if not more strongly predictive of disease severity when measured in the light and support further consideration of evaluating the angle under different lighting conditions.^20^ While the angle is most narrow in the dark on average, it is intuitive that angle configuration in the light could contribute to PACG risk, especially since most people spend the majority of their waking hours in lit environments.

Our study has several limitations. First, we did not separate eyes with PAC and PACG due to the relatively small number of PACG cases in the CHES cohort. Thus, differences in clustering patterns between these two sub-groups are unclear. Second, we analyzed biometric measurements averaged across 8 sectors of the eye. While this approach better captures anatomical variations of the angle, it may weaken the effect of specific sectors that are more predictive of disease severity or progression, such as the temporal and nasal sectors.^32,33^ A more thorough investigation of these factors in the future may help elucidate differences between the proportion of PAC/G in the light and dark. Finally, CHES participants all self-identified as Chinese American. Therefore, the study results may not generalize to other populations. However, generalizability may be improved compared to prior community and hospital based studies.^16–19^

In conclusion, hierarchical cluster analysis appears to classify angle closure eyes by disease severity when applied to biometric measurements that comprise the full spectrum of PACD. In addition, measurements obtained in the light may provide useful information about disease severity even though clinical assessments of the anterior chamber angle are by convention performed in the dark. While clustering of biometric measurements obtained in the light may provide a novel approach to risk-stratify PACD eyes for more severe disease, longitudinal studies using quantitative OCT measurements to predict disease outcomes are needed to elucidate the long-term clinical significance of this approach for identifying patients at higher risk for PACG.

## Data Availability

All data produced in the present study are available upon reasonable request to the authors.

## Table and Figure Captions

**Supplementary Figure 1:** Dendrogram representation of agglomerative hierarchical cluster analysis of PACD eyes in the dark (left) and light (right).

**Supplementary Table 1**. Comparison of demographics and ocular biometric factors between Clusters 1A, 1B, and 2 from the 3-cluster sensitivity analysis

