## Supplementary Table 1 for "Classification of Primary Angle Closure Disease by Hierarchical Cluster Analysis of Ocular Biometrics in the Dark and Light"

| **Supplementary Table 1.**  Comparison of demographics and ocular biometric factors between Clusters 1A, 1B, and 2 from the 3-cluster sensitivity analysis | | | | | | | | | | |
| --- | --- | --- | --- | --- | --- | --- | --- | --- | --- | --- |
| **Lighting** | **Mean Parameters (Mean ± SD)** | **Cluster 1A** |  | **Cluster 1B** |  | **Cluster 2** | **P-value^a^** | **1A v 1B^a^** | **1A v 2^a^** | **1B v 2^a^** |
| Dark |  | N = 71 |  | N = 61 |  | N = 27 |  |  |  |  |
|  | IOP^b^ (mmHg) | 16.638 ± 4.019 |  | 16.054 ± 3.085 |  | 15.321 ± 2.688 | 0.23 | > 0.99 | 0.29 | > 0.99 |
|  | AOD750 (mm) | 0.132 ± 0.044 |  | 0.185 ± 0.044 |  | 0.225 ± 0.055 | **< 0.001** | **< 0.001** | **< 0.001** | **0.01** |
|  | TISA750 (mm^2^) | 0.066 ± 0.024 |  | 0.113 ± 0.027 |  | 0.114 ± 0.024 | **< 0.001** | **< 0.001** | **< 0.001** | 0.1 |
|  | IA (mm^2^) | 1.650 ± 0.249 |  | 1.567 ± 0.153 |  | 1.554 ± 0.213 | 0.10 | 0.11 | 0.12 | > 0.99 |
|  | IT750 (mm) | 0.426 ± 0.085 |  | 0.366 ± 0.059 |  | 0.425 ± 0.052 | **< 0.001** | **< 0.001** | > 0.99 | **< 0.001** |
|  | IC (mm) | 0.270 ± 0.056 |  | 0.330 ± 0.047 |  | 0.206 ± 0.060 | **< 0.001** | **< 0.001** | **< 0.001** | **< 0.001** |
|  | ACD (mm) | 2.160 ± 0.212 |  | 2.109 ± 0.170 |  | 2.420 ± 0.184 | **< 0.001** | 0.39 | **< 0.001** | **< 0.001** |
|  | LV (mm) | 0.800 ± 0.168 |  | 0.882 ± 0.130 |  | 0.573 ± 0.136 | **< 0.001** | **0.006** | > 0.99 | **< 0.001** |
|  | ACW (mm) | 11.524 ± 0.456 |  | 11.492 ± 0.307 |  | 11.454 ± 0.281 | 0.70 | > 0.99 | > 0.99 | > 0.99 |
|  | PD (mm) | 3.848 ± 0.722 |  | 3.463 ± 0.503 |  | 4.259 ± 0.880 | **< 0.001** | **0.001** | 0.13 | **< 0.001** |
| Light |  | N = 110 |  | N = 16 |  | N = 33 |  |  |  |  |
|  | IOP^b^ (mmHg) | 16.015 ± 3.088 |  | 18.312 ± 5.591 |  | 15.747 ± 3.236 | **0.03** | **< 0.001** | > 0.99 | **< 0.001** |
|  | AOD750 (mm) | 0.225 ± 0.050 |  | 0.169 ± 0.061 |  | 0.311 ± 0.047 | **< 0.001** | **< 0.001** | **< 0.001** | **< 0.001** |
|  | TISA750 (mm^2^) | 0.132 ± 0.030 |  | 0.084 ± 0.034 |  | 0.164 ± 0.026 | **< 0.001** | **< 0.001** | **< 0.001** | **< 0.001** |
|  | IA (mm^2^) | 1.848 ± 0.200 |  | 1.551 ± 0.187 |  | 1.888 ± 0.218 | **< 0.001** | **< 0.001** | 0.99 | **< 0.001** |
|  | IT750 (mm) | 0.333 ± 0.065 |  | 0.378 ± 0.051 |  | 0.332 ± 0.046 | **0.01** | **0.003** | > 0.99 | **0.009** |
|  | IC (mm) | 0.314 ± 0.065 |  | 0.212 ± 0.081 |  | 0.256 ± 0.056 | **< 0.001** | **< .001** | **< .001** | **0.22** |
|  | ACD (mm) | 2.119 ± 0.147 |  | 1.977 ± 0.240 |  | 2.464 ± 0.142 | **< 0.001** | **0.003** | **< 0.001** | **< 0.001** |
|  | LV (mm) | 0.862 ± 0.152 |  | 0.898 ± 0.200 |  | 0.656 ± 0.124 | **< 0.001** | **> 0.99** | **< 0.001** | **< 0.001** |
|  | ACW (mm) | 11.520 ± 0.331 |  | 11.235 ± 0.550 |  | 11.685 ± 0.362 | **< 0.001** | **0.01** | 0.07 | **< 0.001** |
|  | PD (mm) | 2.431 ± 0.416 |  | 3.398 ± 0.893 |  | 2.567 ± 0.351 | **< 0.001** | **< 0.001** | 0.11 | **0.003** |
| ACD: Anterior chamber depth; ACW: Anterior chamber width; AOD: Angle opening distance; IA: Iris area; IC: Iris curvature; IOP: Intraocular pressure; IT: Iris thickness; LV: Lens vault; PAC: Primary angle closure; PACS: Primary angle closure suspect; PACG: Primary angle closure glaucoma; PD: Pupillary diameter; TISA: Trabecular iris surface area  ^a^ Statistical significance tested by ANOVA/Pairwise t-test or Kruskal-Wallis/Dunn test  ^b^ Post hoc; not included in cluster analysis | | | | | | | | | | |
