## Supplementary Figure 1 for "Classification of Primary Angle Closure Disease by Hierarchical Cluster Analysis of Ocular Biometrics in the Dark and Light"

**Supplementary Figure 1:** Dendrogram representation of agglomerative hierarchical cluster analysis of PACD eyes in the dark (left) and light (right).

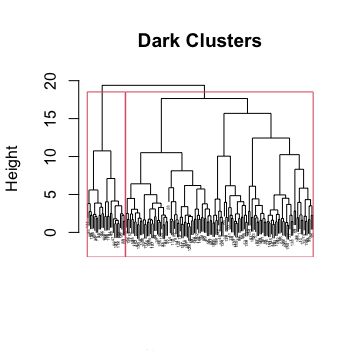

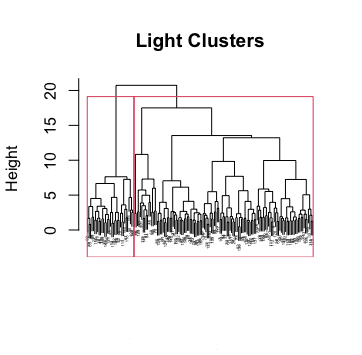
